# Social disparities in flood exposure and associations with the urban environment in 44,698 neighborhoods in 276 cities in eight Latin American countries

**DOI:** 10.1101/2024.07.02.24309839

**Authors:** Josiah L. Kephart, Usama Bilal, Nelson Gouveia, Olga Lucia Sarmiento, Emily Shingara, Karla Rangel Moreno, Maryia Bakhtsiyarava, Juan Pablo Rodriguez, Salvador Ayala, Gabriel Carrasco-Escobar, Ana V Diez Roux, SALURBAL-Climate Project

## Abstract

**Background:** Climate change is expected to greatly increase exposure to flooding, particularly in urban populations in low- and middle-income countries. We examined within-city social disparities in exposure to flooding in 276 Latin American cities and associated features of the neighborhood urban environment.

**Methods:** We used a spatially granular dataset of historical flood events from 2000 to 2018 to describe neighborhood flooding within cities across eight Latin American countries (Argentina, Brazil, Chile, Colombia, Costa Rica, Guatemala, Mexico, and Panama). We estimated the percentage of the population living in flooded neighborhoods, described social disparities in flooding based on neighborhood educational attainment, and compared the magnitude of disparities across and within cities. We used multilevel models to examine how city- and neighborhood-level factors are related to neighborhood flooding.

**Results:** We examined 44,698 neighborhoods in 276 cities from eight countries with a total of 223 million residents and 117 distinct flood events from 2000-2018. One in four residents in neighborhoods in the lowest education quintile lived in neighborhoods with flooding, compared to one in 20 residents of the highest neighborhood education quintile. Greater neighborhood flooding was associated with lower neighborhood-level educational attainment and with neighborhoods that were coastal, less dense (population or intersection), further from the city center, greener, and had steeper slopes. There was no association between city-level educational attainment and flooding.

**Conclusion:** There are large social disparities in neighborhood flooding within Latin American cities. Residents of areas with lower education attainment face substantially higher risks of flooding. Policymakers must prioritize flood adaptation and recovery efforts in neighborhoods with lower socioeconomic position.

## 1. Introduction

Floods are the most frequently occurring climate-related disaster, affecting more people than any other disaster type.^1^ Climate change is projected to increase the frequency, intensity, and extent of major flood events through rising sea levels, faster snowmelt, and a greater frequency of storms with extreme precipitation.^2–4^ Increasing levels of urbanization have exacerbated the risk of flooding in densely populated areas by altering or eliminating natural rainwater sinks and watersheds.^5^ The combined forces of climate change and urbanization have led to rapid growth in the proportion of the global population that live in flood-prone areas.^4,5^ Between 2000 and 2015, the global population affected by floods grew by 58-86 million, an alarming increase of 20-24% within just 16 years.^4^

Increases in population exposure to floods are unequally distributed between countries and are projected to disproportionately impact residents of low- and middle-income countries.^4,6^ Within high-income countries, marginalized communities have greater risk of flood exposure^7–9^ and greater susceptibilities to the health, social, and economic impacts of floods.^9,10^ However, little is known about how flood risk is distributed along social gradients in highly urbanized Latin America, where wide social, health, and environmental inequalities are common^11^ and patterns of neighborhood segregation and environmental disparities often differ from those observed in high-income countries.^12–14^ Furthermore, little is known about what features of the urban environment are associated with differences in flood risk between and within the cities of Latin America. This lack of knowledge on the interplay between floods, population risk, social disparities, and the urban environment is a barrier to climate adaptation planning across Latin America, where 80% of the population lives in highly unequal urban areas^15^ and climate-related flooding is expected to increase in the coming decades.^2^ There is a critical need to examine the linkages between population flood risk, social disparities, and the urban environment to inform flood adaptation efforts that protect public health and decrease inequality across urban Latin America.

To address these gaps, we described social disparities in exposure to recent floods and urban characteristics associated with flooding at the neighborhood level within 276 Latin American cities diverse in climate, socioeconomic factors, and features of the urban environment.

## 2. Results

### 2.1. Study neighborhoods, population, and urban characteristics

We examined 44,698 neighborhoods in 276 cities with recorded flood events in Argentina, Brazil, Chile, Colombia, Costa Rica, Guatemala, Mexico, and Panama (**Table 1, Supplemental Table S1**). The geographic locations of observed cities are presented in **Figure 1**, where study cities (all with recorded flood events) are represented by blue points while excluded cities without recorded flood events (N=50) are indicated as dark gray points. Neighborhoods in Brazil had the lowest median population educational attainment (67.6% of adults aged 25+ completed primary school, interquartile range [IQR] 14.9) while neighborhoods in Mexico (91.8% [IQR 8.7]) had the highest median educational attainment. Neighborhoods in Colombia had the highest population density (11.0 [15.8] thousand residents per km^2^) while neighborhoods in Brazil were the least dense (4.1 [7.5]). Neighborhood and city population and characteristics are described overall and by country in **Table 1**.

**Figure 1:**
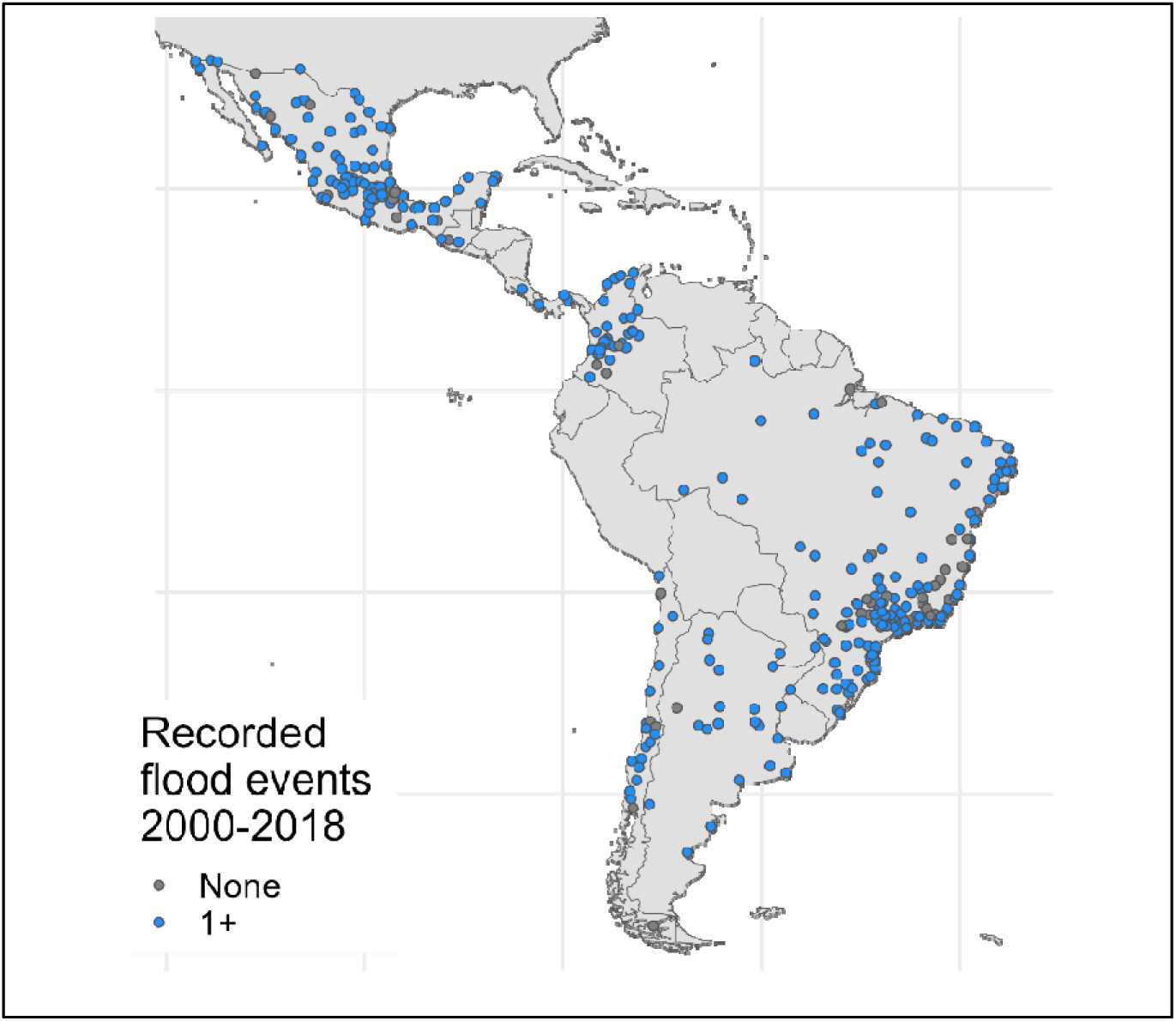
Study cities with recorded floods from 2000-2018. 276 cities had recorded floods and were included in the analysis (blue color), 50 cities had no recorded floods and were excluded (dark gray color).

**Table 1.** Population and urban environment characteristics of 44,698 study neighborhoods in 276 Latin American cities with recorded flood events from 2000-2018, overall and by country. Results presented as number (N) and percent or mean (standard deviation).

|  | **Total** | **Argentina** | **Brazil** | **Central America<sup>a</sup>** | **Chile** | **Colombia** | **Mexico** |
| --- | --- | --- | --- | --- | --- | --- | --- |
| **Neighborhood level** |  |  |  |  |  |  |  |
| N of neighborhoods | 44,698 | 2,002 | 3,660 | 4,929 | 1,824 | 2,907 | 29,376 |
| Population density<sup>b</sup> | 8.1 (11.0) | 8.7 (9.8) | 6.3 (7.1) | 11.1 (24.8) | 5.9 (5.0) | 12.9 (11.9) | 7.5 (6.9) |
| Educational attainment (% primary school or less) | 86.3 (13.6) | 89.4 (7.9) | 67.9 (11.8) | 73.9 (22.0) | 88.9 (9.5) | 88.7 (14.5) | 90.5 (8.0) |
| Intersection density<sup>c</sup> | 150 (172) | 81 (43) | 89 (57) | 101 (112) | 135 (101) | 251 (544) | 162 (100) |
| Greenness (NDVI)<sup>d</sup> | 0.47 (0.20) | 0.46 (0.20) | 0.57 (0.19) | 0.60 (0.19) | 0.46 (0.22) | 0.57 (0.20) | 0.42 (0.18) |
| Distance from city center (km) | 12.7 (13.8) | 21.2 (19.2) | 14.9 (14.5) | 11.8 (8.1) | 9.2 (12.0) | 9.7 (12.4) | 12.5 (14.1) |
| N (%) of coastal neighborhoods | 2,244 (5.0%) | 149 (7.4%) | 592 (16.2%) | 160 (3.2%) | 358 (19.6%) | 182 (6.3%) | 803 (2.7%) |
| Altitude (meters) | 1,063 (921) | 134 (287) | 409 (369) | 1,409 (675) | 218 (429) | 1,163 (992) | 1,253 (943) |
| Slope (degrees) | 4.4 (3.8) | 2.8 (2.0) | 5.2 (3.4) | 7.0 (5.3) | 5.0 (4.4) | 6.0 (5.7) | 3.8 (3.0) |
| **City level** |  |  |  |  |  |  |  |
| N of cities | 276 | 21 | 124 | 5 | 16 | 29 | 81 |
| City population (thousands) | 721 (2,036) | 1,023 (3,078) | 715 (2,021) | 1,130 (1,018) | 228 (158) | 759 (1,479) | 711 (2,168) |
| GDP per capita (USD \$1000s) | 15.5 (11.3) | 17.4 (7.2) | 15.2 (8.1) | 16.9 (7.4) | 21.2 (18.3) | 10.3 (3.8) | 16.1 (15.5) |
| Population density<sup>b</sup> | 7.1 (3.8) | 5.6 (1.3) | 6.1 (2.1) | 7.5 (2.7) | 7.0 (1.4) | 15.7 (5.1) | 5.9 (1.5) |
| Education (% primary school or less) | 74.1 (10.0) | 79.6 (2.8) | 65.6 (6.1) | 86.6 (6.1) | 89.5 (2.5) | 83.9 (3.7) | 78.2 (7.0) |
| Intersection density<sup>c</sup> | 92.3 (25.3) | 85.1 (16.3) | 84.4(19.2) | 60.5 (23.7) | 125.1 (15.1) | 117.6 (31.1) | 92.9 (23.4) |
| Greenness (NDVI) | 0.56 (0.12) | 0.52 (0.11) | 0.60 (0.06) | 0.69 (0.06) | 0.42 (0.20) | 0.63 (0.07) | 0.50 (0.14) |

<sup>a</sup>Central America grouping includes urban neighborhoods in Costa Rica (N=1 city), Guatemala (N=1), and Panama (N=3)
<sup>b</sup>Thousands of residents per square kilometer
<sup>c</sup>Street intersections per square kilometer
<sup>d</sup>Median normalized vegetation index within the neighborhood or city

### 2.2. Neighborhood flood population exposures and educational disparities

There were 117 unique floods across the 276 study cities between 2000 and 2018. Large-scale floods often affected more than one city or country. The number of flood events ranged from 12 floods in Chile to 62 floods in Brazil (**Table 2**). **Supplemental Table 2** shows the number of flood events by individual city. Of the 228.3 million residents residing in study cities as of the last census, 38.1 million people (16.7% of residents) lived in neighborhoods that flooded at least once between 2000 and 2018 (**Table 2**). **Supplemental Figure S2** shows neighborhood flooding within two select cities: the metropolitan area of Buenos Aires, Argentina and Medellín, Colombia. We observed large differences by country in the percentage of urban residents in neighborhoods with flooding. While 24.0% of residents of Brazilian cities and 19.5% of residents of Colombian cities lived in neighborhoods with flooding, just 4.3% of residents in Central American cities lived in flooded neighborhoods. (**Table 2**).

**Table 2.** Neighborhood and population exposures to flood events, by neighborhood education attainment and country in 44,698 neighborhoods in 276 cities in eight Latin American countries from 2000-2018.

|  | Total,<br>neighborhoods | Number of unique<br>flood events** | City-specific % of<br>neighborhoods<br>with flooding:<br>median (P10, P90) | Total<br>Population<br>(millions) | Population in<br>flooded<br>neighborhoods<br>(millions) | % of total<br>population in<br>flooded<br>neighborhoods |
| --- | --- | --- | --- | --- | --- | --- |
| **Overall** | 44,698 | 117 | 14% (1, 56) | 228.3 | 38.1 | 16.7% |
| **By country grouping** |  |  |  |  |  |  |
| Argentina | 2,002 | 43 | 31% (7, 66) | 23.2 | 2.8 | 12.1 % |
| Brazil | 3,660 | 62 | 23% (8, 75) | 101.0 | 24.4 | 24.0 % |
| Central American* | 4,929 | 17 | 2% (1, 6) | 6.2 | 0.3 | 4.3 % |
| Chile | 1,824 | 12 | 5% (2, 26) | 5.1 | 0.4 | 8.2 % |
| Colombia | 2,907 | 23 | 17% (3, 37) | 22.4 | 4.4 | 19.5 % |
| Mexico | 29,376 | 31 | 2% (1, 11) | 70.0 | 5.9 | 8.4 % |
| **By neighborhood**<br>**educational attainment** |  |  |  |  |  |  |
| 1<sup>st</sup> quintile (lowest) | 8,940 | - | - | 97.6 | 23.5 | 24.0 % |
| 2<sup>nd</sup> | 8,940 | - | - | 42.1 | 7.6 | 18.2 % |
| 3<sup>rd</sup> | 8,940 | - | - | 34.4 | 3.1 | 9.1 % |
| 4<sup>th</sup> | 8,939 | - | - | 32.7 | 2.7 | 8.2 % |
| 5<sup>th</sup> quintile (highest) | 8,939 | - | - | 21.5 | 1.2 | 5.6 % |

\*Includes cities in the countries of Costa Rica (N=1 city), Guatemala (N=1), and Panama (N=3).
\*\*The number of unique flood events overall does not equal the sum of unique flood events by country because some floods affected multiple countries (as well as multiple cities).

We also observed large dose-response disparities in flooding by neighborhood educational attainment. Across the distribution of 44,698 study neighborhoods, 24.0% of residents of neighborhoods in the lowest quintile of education attainment lived in flooded neighborhoods, while only 5.6% of residents of the highest educational quintile neighborhoods lived in neighborhoods with flooding (**Table 2**, **Figure 2**). Across study cities, residents of the lowest education neighborhoods were 4.3 times more likely to live in neighborhoods with flooding than residents of the highest education neighborhoods. We also examined the relationship between neighborhood education and two or more neighborhood floods. The association of higher neighborhood education with lower probability of neighborhood flooding was consistent across all education quintiles and when comparing single (**Table 2**) and multiple floods (**Figure 2**).

**Figure 2:**
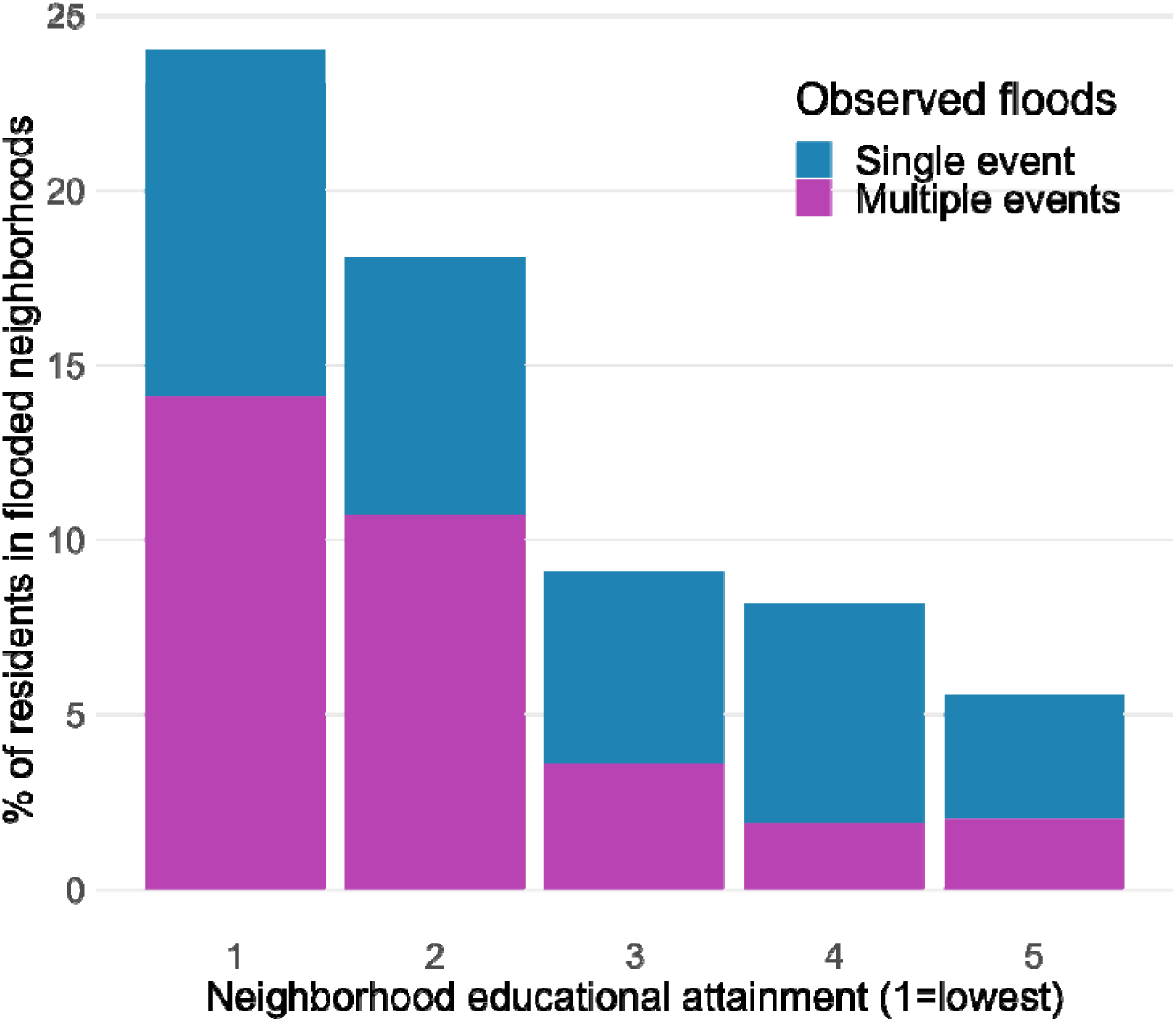
Percent of total residents experiencing flooding in their neighborhoods in 276 Latin American cities with recorded flood events between 2000-2018, by neighborhood education attainment quintile.

When we examined within-city disparities in flooding in each of 276 cities, disparities by neighborhood education were even more pronounced (**Table 3**). Across all 276 cities, 80.1% of cities showed educational disparities in flooding with higher prevalence of flooding in neighborhoods of lower education. The median slope index of inequality (SII) for flooding was 8.3, indicating that the prevalence of flooding was 8.3 times higher in neighborhoods of the lowest vs highest educational quintile. These disparities were very heterogenous, with 25% of cities having an SII at or below 1.3 (i.e. prevalence of flooding was 1.3 times higher in neighborhoods of the lowest vs highest educational quintile) and 25% of cities having an SII at or above 100 (i.e. prevalence of flooding was 100 times higher in neighborhoods of the lowest vs highest educational quintile). This overall pattern of substantially higher odds of flooding in lowest vs. highest education neighborhoods within cities was consistent across all countries, although the median SII for Mexican cities was substantially higher than for other countries (25.4 vs 3.0-8.6). In many cities, the RR was greater than 100, indicating that floods occurred almost exclusively in lower education neighborhoods. When stratifying cities by size (quintile of population), we found more extreme results in smaller cities.

**Table 3.** Within-city disparities in flood exposure by neighborhood educational attainment within 276 cities in eight countries of Latin America that experienced flooding between 2000-2018. Shown are city-specific slope indices of inequality of flooding in neighborhoods comparing lowest vs. highest education quintiles in each city.

|  | City-specific slope indices of inequality of flooding in lowest vs. highest education neighborhoods |  |  | % of cities with higher risk of flooding in lowest vs. highest education quintile neighborhoods (SII>1) |
| --- | --- | --- | --- | --- |
|  | 25<sup>th</sup> percentile | Median | 75<sup>th</sup> percentile |  |
| **Overall** | 1.3 | 8.3 | > 100 | 80.1% |
| **By country grouping** |  |  |  |  |
| Argentina | 1.6 | 5.6 | 27.5 | 85.7% |
| Brazil | 1.0 | 4.0 | > 100 | 73.4% |
| Central American* | 2.0 | 5.0 | 9.4 | 80.0% |
| Chile | 1.8 | 8.6 | > 100 | 81.2% |
| Colombia | 1.7 | 3.0 | 30.9 | 86.2% |
| Mexico | 2.3 | 25.4 | > 100 | 86.4% |
| **By city population size** |  |  |  |  |
| 1<sup>st</sup> quintile (smallest) | 2.5 | 25.2 | >100 | 82.1% |
| 2<sup>nd</sup> | 1.2 | 8.7 | >100 | 78.2% |
| 3<sup>rd</sup> | 2.0 | 9.6 | >100 | 85.5% |
| 4<sup>th</sup> | 1.2 | 13.6 | >100 | 80.0% |
| 5<sup>th</sup> quintile (largest) | 1.0 | 2.13 | 8.2 | 74.5% |

\*Includes cities in the countries of Costa Rica (N=1 city), Guatemala (N=1), and Panama (N=3).

### 2.3. Associations between neighborhood flooding and the urban environment

In **Table 4**, we present associations between neighborhood flooding and features of the urban environment. In multilevel models adjusting for both neighborhood and city characteristics simultaneously, higher neighborhood population density (OR 0.47 per unit higher z-score [95% confidence interval (CI) 0.41 to 0.52]) and intersection density (OR 0.71 per unit higher z-score [95% CI 0.64 to 0.77]) were associated with lower odds of flooding. Coastal neighborhoods were substantially more likely to experience flooding than non-coastal neighborhoods (OR 46.24 [95% CI 38.30, 55.82]). We found that neighborhoods that were greener (OR 1.43 per unit higher z-score [95% CI 1.32, 1.55]) and more distant from the city center (OR 1.19 per unit higher z-score [95% CI 1.13, 1.25]) were more likely to experience flooding compared to less green, central neighborhoods. In this same model, higher altitude and flatter slope were also associated with higher odds of neighborhood flooding. In our sensitivity analysis restricting these models to the 196 cities with two or more flood events, we found nearly identical associations, suggesting that flood patterns within cities are reasonably captured by as few as one recorded flood event (**Supplemental Table S3)**.

**Table 4.**
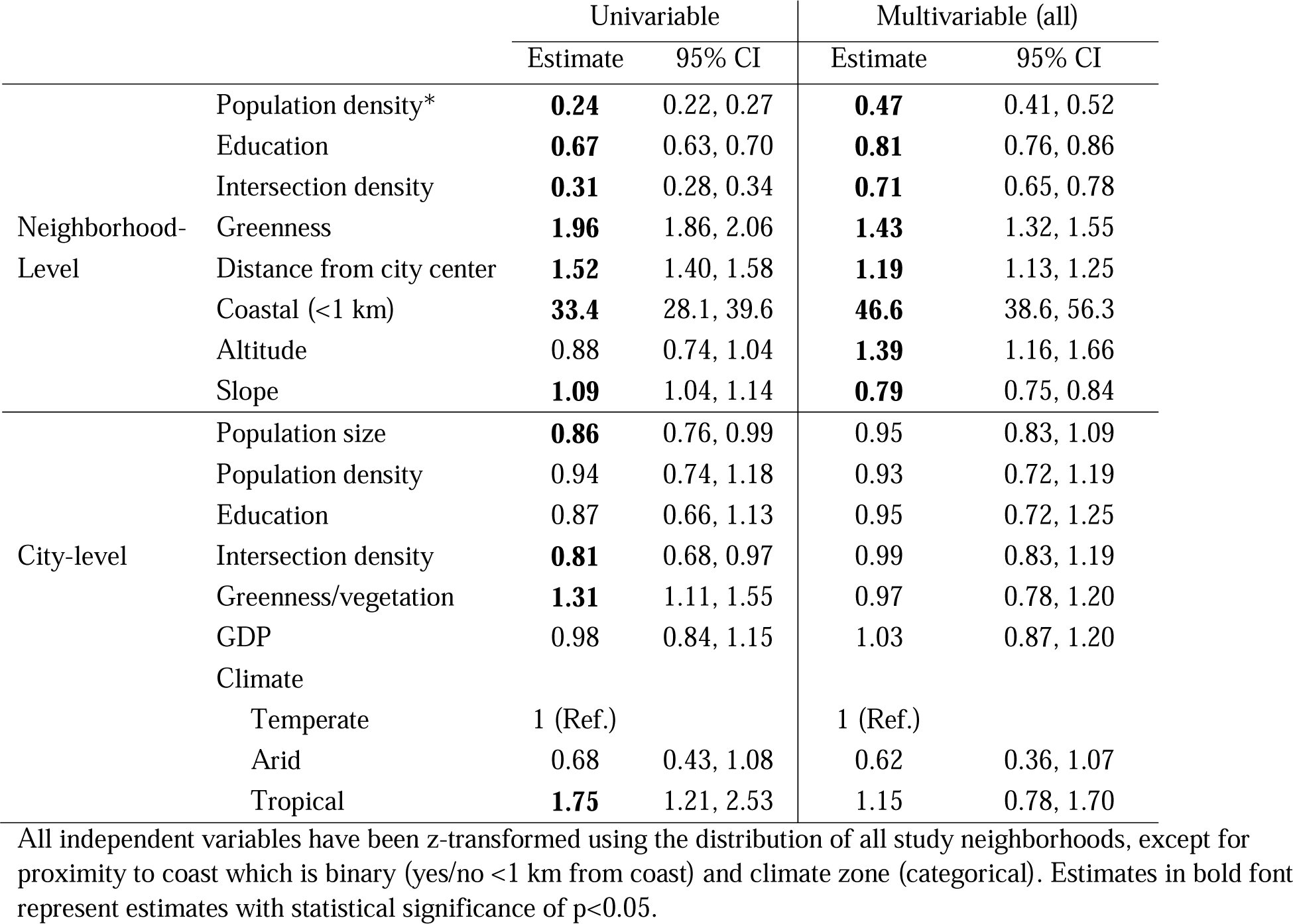
Odds ratios of neighborhood flooding associated with a one standard deviation higher value for neighborhood- and city-level features in 44,698 neighborhoods in 276 cities in eight Latin American countries from 2000-2018.

|  |  | Univariable |  | Multivariable (all) |  |
| --- | --- | --- | --- | --- | --- |
|  |  | Estimate | 95% CI | Estimate | 95% CI |
| Neighborhood-Level | Population density* | **0.24** | 0.22, 0.27 | **0.47** | 0.41, 0.52 |
|  | Education | **0.67** | 0.63, 0.70 | **0.81** | 0.76, 0.86 |
|  | Intersection density | **0.31** | 0.28, 0.34 | **0.71** | 0.65, 0.78 |
|  | Greenness | **1.96** | 1.86, 2.06 | **1.43** | 1.32, 1.55 |
|  | Distance from city center | **1.52** | 1.40, 1.58 | **1.19** | 1.13, 1.25 |
|  | Coastal (<1 km) | **33.4** | 28.1, 39.6 | **46.6** | 38.6, 56.3 |
|  | Altitude | 0.88 | 0.74, 1.04 | **1.39** | 1.16, 1.66 |
|  | Slope | **1.09** | 1.04, 1.14 | **0.79** | 0.75, 0.84 |
| City-level | Population size | **0.86** | 0.76, 0.99 | 0.95 | 0.83, 1.09 |
|  | Population density | 0.94 | 0.74, 1.18 | 0.93 | 0.72, 1.19 |
|  | Education | 0.87 | 0.66, 1.13 | 0.95 | 0.72, 1.25 |
|  | Intersection density | **0.81** | 0.68, 0.97 | 0.99 | 0.83, 1.19 |
|  | Greenness/vegetation | **1.31** | 1.11, 1.55 | 0.97 | 0.78, 1.20 |
|  | GDP | 0.98 | 0.84, 1.15 | 1.03 | 0.87, 1.20 |
|  | Climate |  |  |  |  |
|  | Temperate | 1 (Ref.) |  | 1 (Ref.) |  |
|  | Arid | 0.68 | 0.43, 1.08 | 0.62 | 0.36, 1.07 |
|  | Tropical | **1.75** | 1.21, 2.53 | 1.15 | 0.78, 1.70 |

All independent variables have been z-transformed using the distribution of all study neighborhoods, except for proximity to coast which is binary (yes/no <1 km from coast) and climate zone (categorical). Estimates in bold font represent estimates with statistical significance of $p < 0.05$ .

Our descriptive findings of educational disparities in neighborhood flooding were consistent even after adjusting for urban features. After simultaneously adjusting for a suite of city and neighborhood-level urban and natural features, neighborhoods with higher education had lower odds of flooding than neighborhoods with lower education (OR 0.81 per unit higher educational attainment z-score [95% confidence interval (CI) 0.75 to 0.86]). However, we did not find an association between city-level education and flooding risk.

## 3. Discussion

We conducted a descriptive analysis at high spatial resolution of flooding across nearly 45,000 neighborhoods of 276 cities in eight Latin American countries, representing 228 million residents. Our study provides four key findings. First, 17% of the 228 million residents of these cities lived in neighborhoods with at least one flood during the study period. Second, there are significant disparities in flood exposure by education attainment across Latin American cities. Across the region, residents of the lowest education quintile neighborhoods were 4.3 times more likely to live in a neighborhood that had flooded than residents of highest education quintile neighborhoods. Third, there are massive disparities within cities of which neighborhoods do and do not experience flooding. Within each of 276 cities, the median risk of neighborhood flooding was eight times higher in neighborhoods with the lowest education levels than those with the highest education levels within the same city. Overall, 80% of cities demonstrated educational inequities in flood exposure, a pattern consistent across all study countries. Fourth, among Latin American cities, coastal neighborhoods and neighborhoods that are less dense, greener, and peripheral to the city center were more likely to flood. These findings emphasize the need for socially informed climate adaptation policies that protect marginalized neighborhoods where the risk of flooding may be greatest.

Our study identified consistent patterns of social inequities in neighborhood flooding within cities across Latin America. We are unaware of other studies that report social disparities in floods in Latin America at the neighborhood level, yet our findings corroborate other studies at higher geographic levels. Rentschler et al. examined flood exposure and poverty in 188 countries, with some subnational analyses at the department or state level, finding that 89% of the world’s flood-exposed people live in low - and middle-income countries.^6^ Within Latin America, Rasch et al. conducted a national analysis of Brazil at the municipal level, finding that differences in income across municipalities significantly predicts risk of flood hazards.^16^ While there is a growing recognition of environmental injustices in flood exposure at the national and subnational levels^7^, we provide novel evidence of detrimental social inequities in neighborhood flooding within cities in Latin America. In the median city, we found that lowest education neighborhoods have eight times greater risk of flooding than the highest education neighborhoods and that these inequities persist (but vary) across all study countries. Additionally, we found that neighborhoods with lower density (population and intersection), more vegetation, and that are peripheral to the urban core face a higher risk of flooding. This may reflect patterns of social segregation and urban infrastructure in Latin America, where in many cities, communities on the urban periphery are more likely to experience lower socioeconomic status^12^ and inferior municipal services.

Our study should be interpreted in the context of some limitations. Although we examined neighborhood flooding at an exceptionally fine spatial scale, we were unable to examine or draw conclusions regarding personal exposure to a flood event. Nevertheless, flooding is directly and indirectly disruptive to communities even beyond injury to specific individual people or houses. Furthermore, while the GFD provides unprecedented, high spatial resolution estimates of flooding events, particularly for low- and middle-income countries, it represents 29% of floods events reported in the DFO and our findings should not be interpreted as estimates of the prevalence of flooding in Latin America cities. For this reason, we emphasize within-city variation among the subset of cities with recorded floods in the GFD. As a sensitivity analysis we included only cities with at least two recorded floods and found similar findings, suggesting that within-city patterns of social disparities in flood exposure are somewhat consistent across flood events. Additionally, we found an association between higher odds of flooding and higher greenness (NDVI) in models adjusted for population and intersection density, slope, altitude, and distance from city center. This may reflect higher levels of natural vegetation in flood plains, wetlands, and other flood-prone areas as greater concentration of water correlates with both flood risk and vegetation. We were unable to include other indicators of the ecological and geological capacity for water accumulation or for density of rivers and tributaries. Further research which incorporates additional ecological information and longitudinal analyses may help shed light on the potentially bidirectional linkages between vegetation and neighborhood flood risk, and how these linkages vary by climate.

Our findings reinforce the importance of prioritizing the needs of socially marginalized communities in environmental risk management and climate adaptation policies. We found significant within-city disparities in flood exposure and higher flood risk among peripheral communities, underscoring the need for urban policymakers to allocate resources towards mitigating the human impacts of flooding and not only economic impacts. While environmental inequities between the global north and global south are substantial, these disparities can also be found at a local level.

Our analysis integrated high-quality satellite-derived estimates of large, historical flood events from 2000-2018 with harmonized census records from eight countries to examine social disparities in flood exposure across 228 million residents of 44,698 neighborhoods in 276 Latin American cities. We found consistent intra-urban disparities in flood exposure, with less educated, coastal, and peripheral communities often more vulnerable. These patterns persisted across all study countries. As climate change increases the risks of human exposure to floods, our findings highlight the urgent need for policymakers to implement environmental and climate adaptation actions that protect marginalized communities and promote environmental and health equity.

## 2. Methods

### 2.1. Study setting

We conducted this study as part of the *Salud Urbana en América Latina* (SALURBAL) project. This international scientific collaboration has compiled and harmonized data on social, environmental, and health characteristics for all cities of 100,000 residents or more in 2010 in 11 Latin American countries (n=371)^17^. In SALURBAL, cities are defined as clusters of administrative units (i.e., municipalities) or single administrative units encompassing the visually apparent built-up area of urban agglomerations as identified using satellite imagery^18^and include a diverse set of cities, from small cities to megacities.

We excluded cities in El Salvador, Nicaragua, and Peru due to the lack of available data at the neighborhood level. Because this study focuses on within-city disparities in flood risk, we excluded 50 cities without recorded flood events within city boundaries. Accordingly, we examined neighborhoods in 276 cities in Argentina, Brazil, Chile, Colombia, Costa Rica, Guatemala, Mexico, and Panama. Neighborhood administrative units varied in name and official definition by country. We used the country-specific, small-area administrative units most analogous to U.S. census tracts, henceforth referred to as “neighborhoods.” Across countries, these neighborhoods had a median population of 2,137 and a median area of 0.36 km^2^ (equivalent to a square with 0.6 km sides). Detailed information on the administrative units used for each country is available in **Supplemental Table S1**.

### 2.2. Neighborhood flood exposures

Estimates of recent flooding were extracted from the Global Flood Database (GFD).^4^ The GFD examines flood events identified by the Dartmouth Flood Observatory (DFO). The DFO monitors and compiles generally large-media-coverage flood events from news sources from every country in the world, synthesizing information from news reports to estimate flood start and end dates and the geographical area affected. For floods identified in the DFO database, the GFD team analyzed satellite imagery to estimate the extent and duration of individual flood events at daily, 250 meter resolution extending the geographic area of interest to include all watersheds intersecting with DFO records of flood-affected areas. A subset (29%) of global DFO events where analyses of satellite data provided useful, high quality estimates of the spatial extent of flooding were included in the GFD. The primary reasons for flood events in the DFO not being included in the GFD database were persistent cloud cover or a lack of detectable inundation at a 250 m resolution within the geographic areas reported as flood-affected by news reports (as compiled by DFO). The final GFD product is a database of global raster datasets showing the maximum spatial extent of flooding at 250 m resolution for 913 large flood events worldwide from 2000 to 2018.^4^ Further details on the GFD data sources and methods have been previously published.^4^ To link flood frequency data to city and neighborhood data, we overlayed floods events in the GFD with SALURBAL city and neighborhood boundaries. We considered a neighborhood to have flooded if flooding occurred anywhere within the neighborhood boundary. The resulting analysis dataset was the cumulative number of recorded, distinct flood events between 2000-2018 for each neighborhood in 276 Latin American cities.

### 2.3. Neighborhood and city characteristics

We used data on neighborhood characteristics and population compiled from national census bureaus and other sources by the SALURBAL project^18^. We used the most recent available census for each country with census years ranging from 2002 to 2018. Information on the year of each census used is available in **Supplemental Table S1**. We compiled neighborhood and city educational attainment (% of the population aged 25 years or older who completed primary education or above) from censuses, as this was the most relevant socioeconomic variable available across all eight national census surveys. At both the neighborhood and city levels, the SALURBAL project previously estimated population density (population divided by built-up area)^18^, intersection density (density of the set of nodes with more than one street emanating from them per km^2^ of built-up area)^18^, and area median greenness measured by the normalized difference vegetation index (NDVI)^19^. We defined neighborhoods as coastal if their spatial boundaries were within 1 km of an ocean coastline. For each neighborhood, we calculated distance from the city center as the Euclidean distance (km) between the neighborhood centroid and, generally, city hall. Neighborhood mean elevation and mean slope were estimated using a well-established 30m resolution topography model^20^, excluding permanent water. At the city level, we also used population estimates from national censuses and statistical agencies^11^, GDP per capita (computed as purchasing power parities in constant 2011 international USD of each city in 2015 derived from regional estimates)^21^, and Köppen climate zone (1^st^ level)^22^. **Appendix I** details the creation of urban environment variables.

### 2.4. Statistical analysis

We calculated summary statistics of neighborhood-level exposures to flooding by country and city and stratified by neighborhood educational attainment overall and by country. We estimated the total population in cities with recorded flooding and the number and percentage of the total city population residing in neighborhoods that had experienced flooding. We grouped neighborhoods into quintiles of neighborhood educational attainment (% of the population aged 25 years or older who completed primary education or above) based on the full sample of study neighborhoods. Due to the small number of cities represented by each country in Central America, we pooled the cities of Costa Rica (N=1 city), Guatemala (N=1), and Panama (N=3) into a single grouping for all analyses.

To describe magnitudes of within-city disparities in flooding, we used a city-specific Poisson model with neighborhood flooding (yes/no) as the outcome and city-specific quintiles of education as the exposure, operationalized as ordinal and rescaled from 0 to 1. This makes the interpretation of the education coefficient analogous to a slope index of inequality (SII): the relative risk (RR) of flooding comparing lowest vs highest levels of education, while considering the entire distribution of education. A city-specific RR of > 1 signifies higher prevalence of flooding in lower education neighborhoods.

To examine associations of city and neighborhood characteristics with flooding, we used multilevel logistic models with a random intercept for each city and fixed effect for country. All neighborhood variables were operationalized as z-scores of the overall distribution of 44,698 neighborhoods for each respective variable, except for coastal neighborhood (yes/no), altitude (meters above sea level), and mean slope (degrees). Similarly, city-level variables were operationalized as z-scores of the overall distribution of 276 cities for each variable, with the exception of climate zone (categorical: temperate, arid, tropical). We first conducted a univariable analysis of each independent variable and the dependent variable of neighborhood flooding. We then assessed all variables for collinearity using Spearman correlation coefficients (**Supplemental Figure S1**). No covariates had correlation coefficients greater than 0.57 (city greenness vs. neighborhood greenness), so we decided to model all neighborhood- and city-level predictors together as a single multivariable model. As a sensitivity analysis, we repeated all models above but leveraged the same multivariable modeling approach but using only the 196 cities with at least two floodings events. Data processing and analyses were conducted in R version 4.3.2^23^.

## Data Availability

Data produced in the present study will be made publicly available after publication.

## Acknowledgments

This study was financially supported by the Wellcome Trust (227810/Z/23/Z). We acknowledge the contribution of all SALURBAL project team members. For more information on SALURBAL and to see a full list of investigators, see https://drexel.edu/lac/salurbal/team/. SALURBAL acknowledges the contributions of many different agencies in generating, processing, facilitating access to data or assisting with other aspects of the project. Please visit https://drexel.edu/lac/data-evidence for a complete list of data sources. JLK was supported by the Drexel FIRST (Faculty Institutional Recruitment for Sustainable Transformation) Program funded by the National Institutes of Health (grant number U54CA267735-02), and the Cotswold Foundation Postdoctoral Fellowship (grant number 230356). UB was also supported by Office of the Director of the National Institutes of Health (award number DP5OD026429).

