## Supplemental Materials for "Social disparities in flood exposure and associations with the urban environment in 44,698 neighborhoods in 276 cities in eight Latin American countries"

### **Supplemental Table S1**: Neighborhood administrative units and years of most recent available census.

| **Country** | **Administrative unit (neighborhood)** | **Census year** |
| --- | --- | --- |
| Argentina | *Fraccion Censal* | 2010 |
| Brazil | *Áreas de Ponderação* | 2010 |
| Chile | *Zona Censal* | 2017 |
| Colombia | *Sector Urbano, Clase = 1* | 2018 |
| Costa Rica | *Distrito* | 2011 |
| Guatemala | *Sector Censal* | 2002 |
| Mexico | *Área Geoestadistica Básica* | 2010 |
| Panama | *Barrio* | 2010 |

**Supplemental Table S2: Observed neighborhood count and population and flood events for 276 study cities.**

| **Country** | **City name** | **Number of neighborhoods observed** | **City population (in census year)** | **Median neighborhood population** | **Flood events observed** |
| --- | --- | --- | --- | --- | --- |
| Argentina | Bahia Blanca | 32 | 301,572 | 8,720 | 2 |
|  | Buenos Aires | 1,256 | 14,531,119 | 10,232 | 9 |
|  | Comodoro Rivadavia | 19 | 186,583 | 10,627 | 1 |
|  | Concordia | 16 | 170,033 | 9,066 | 24 |
|  | Cordoba | 120 | 1,554,755 | 13,054 | 1 |
|  | Corrientes | 22 | 358,180 | 16,625 | 21 |
|  | Formosa | 22 | 234,354 | 7,809 | 24 |
|  | Jujuy | 21 | 317,880 | 16,969 | 5 |
|  | Mar del Plata | 82 | 616,973 | 7,028 | 4 |
|  | Rawson-Trelew | 13 | 131,313 | 12,550 | 1 |
|  | Rio Cuarto | 29 | 246,393 | 8,487 | 7 |
|  | Rosario | 104 | 1,350,855 | 13,052 | 15 |
|  | Salta | 33 | 579,665 | 18,095 | 21 |
|  | San Carlos de Bariloche | 19 | 133,500 | 7,191 | 1 |
|  | San Luis | 19 | 204,019 | 13,182 | 1 |
|  | San Miguel de Tucuman-Tafi Viejo | 66 | 994,553 | 13,591 | 9 |
|  | San Nicolas de los Arroyos | 20 | 136,962 | 6,616 | 15 |
|  | Santa Fe | 35 | 525,093 | 13,284 | 21 |
|  | Santiago del Estero | 34 | 409,404 | 14,190 | 13 |
|  | Tandil | 22 | 123,871 | 4,376 | 2 |
|  | Villa Mercedes | 18 | 125,899 | 5,417 | 4 |
| Brazil | Angra dos Reis | 7 | 169,511 | 19,945 | 2 |
|  | Aracaju | 29 | 835,816 | 28,110 | 2 |
|  | Aracatuba | 11 | 181,579 | 13,996 | 5 |
|  | Araguaina | 7 | 150,484 | 22,438 | 4 |
|  | Araguari | 6 | 109,801 | 18,500 | 3 |
|  | Arapiraca | 12 | 214,006 | 15,554 | 1 |
|  | Araraquara | 17 | 243,140 | 14,588 | 1 |
|  | Araras | 7 | 118,843 | 16,974 | 3 |
|  | Araruama | 10 | 183,907 | 18,813 | 2 |
|  | Atibaia | 8 | 146,311 | 14,386 | 2 |
|  | Balneario Camboriu | 10 | 170,450 | 15,270 | 6 |
|  | Barreiras | 7 | 137,427 | 18,226 | 2 |
|  | Barretos | 6 | 112,101 | 17,976 | 4 |
|  | Belem | 65 | 2,101,883 | 32,621 | 1 |
|  | Belo Horizonte | 174 | 4,707,362 | 26,724 | 2 |
|  | Bento Goncalves | 11 | 163,159 | 13,814 | 1 |
|  | Birigui | 6 | 108,728 | 16,865 | 5 |
|  | Blumenau | 29 | 486,379 | 15,648 | 1 |
|  | Boa Vista | 13 | 263,407 | 18,689 | 5 |
|  | Botucatu | 7 | 127,328 | 17,199 | 5 |
|  | Braganca Paulista | 9 | 146,744 | 14,818 | 4 |
|  | Brasilia | 78 | 3,187,984 | 40,502 | 3 |
|  | Cabo Frio | 14 | 301,817 | 19,032 | 2 |
|  | Campina Grande | 24 | 439,164 | 16,874 | 2 |
|  | Campinas | 114 | 2,783,254 | 22,052 | 5 |
| Brazil | Campo Grande | 8 | 786,797 | 103,857 | 2 |
|  | Campos dos Goytacazes | 9 | 463,731 | 26,514 | 3 |
|  | Caraguatatuba | 10 | 174,782 | 14,628 | 6 |
|  | Caruaru | 12 | 314,912 | 15,682 | 1 |
|  | Cascavel | 17 | 286,205 | 15,861 | 1 |
|  | Caxias | 7 | 155,129 | 17,917 | 1 |
|  | Caxias do Sul | 27 | 435,564 | 14,073 | 4 |
|  | Chapeco | 10 | 158,894 | 16,024 | 14 |
|  | Criciuma | 18 | 318,283 | 16,398 | 8 |
|  | Cuiaba | 35 | 803,694 | 20,735 | 10 |
|  | Curitiba | 100 | 2,857,466 | 28,032 | 11 |
|  | Divinopolis | 7 | 213,016 | 34,904 | 1 |
|  | Dourados | 10 | 196,035 | 18,930 | 8 |
|  | Feira de Santana | 16 | 589,925 | 36,154 | 1 |
|  | Florianopolis | 57 | 851,955 | 13,678 | 8 |
|  | Fortaleza | 99 | 3,411,846 | 32,645 | 3 |
|  | Foz do Iguacu | 11 | 256,088 | 14,910 | 13 |
|  | Garanhuns | 7 | 129,408 | 16,084 | 1 |
|  | Goiania | 69 | 2,013,288 | 29,664 | 2 |
|  | Guarapari | 6 | 105,286 | 16,407 | 1 |
|  | Guarapuava | 9 | 167,328 | 18,607 | 2 |
|  | Ilheus | 9 | 184,236 | 18,385 | 1 |
|  | Imperatriz | 15 | 290,946 | 18,209 | 4 |
|  | Itabira | 5 | 109,783 | 23,020 | 1 |
|  | Itajai | 16 | 308,534 | 16,146 | 7 |
|  | Jaragua do Sul | 11 | 193,611 | 16,029 | 1 |
|  | Jau | 7 | 131,040 | 16,552 | 5 |
|  | Jequie | 7 | 151,895 | 21,372 | 1 |
|  | Ji-Parana | 6 | 116,610 | 16,053 | 5 |
|  | Joao Pessoa | 26 | 1,010,861 | 27,707 | 3 |
|  | Joinville | 30 | 540,098 | 16,847 | 7 |
|  | Juazeiro do Norte | 21 | 426,690 | 17,859 | 2 |
|  | Jundiai | 34 | 596,148 | 15,216 | 3 |
|  | Lages | 9 | 156,727 | 17,793 | 16 |
|  | Limeira | 7 | 296,051 | 43,443 | 3 |
|  | Linhares | 7 | 141,306 | 19,684 | 1 |
|  | Londrina | 13 | 651,632 | 28,202 | 2 |
|  | Macae | 11 | 188,974 | 15,700 | 2 |
|  | Maceio | 28 | 1,061,809 | 32,530 | 3 |
|  | Manaus | 33 | 1,802,014 | 50,359 | 5 |
|  | Maraba | 4 | 233,669 | 57,742 | 1 |
|  | Mogi Guacu | 13 | 233,794 | 16,622 | 1 |
|  | Montes Claros | 22 | 361,915 | 15,740 | 1 |
|  | Mossoro | 13 | 259,815 | 19,107 | 3 |
|  | Natal | 43 | 1,227,675 | 28,899 | 3 |
|  | Ourinhos | 5 | 103,035 | 20,664 | 1 |
|  | Palmas | 10 | 228,332 | 19,111 | 5 |
|  | Paranagua | 7 | 140,469 | 15,886 | 7 |
|  | Parauapebas | 7 | 153,908 | 21,647 | 1 |
|  | Parnaiba | 5 | 145,705 | 27,949 | 3 |
|  | Parobe | 8 | 161,653 | 19,392 | 4 |
| Brazil | Passo Fundo | 10 | 184,826 | 16,248 | 8 |
|  | Passos | 6 | 106,290 | 15,060 | 4 |
|  | Patos de Minas | 9 | 138,710 | 14,087 | 1 |
|  | Pelotas | 21 | 352,573 | 16,052 | 16 |
|  | Petrolina | 25 | 491,927 | 18,274 | 4 |
|  | Piracicaba | 20 | 364,571 | 18,116 | 5 |
|  | Pocos de Caldas | 9 | 152,435 | 13,738 | 1 |
|  | Ponta Grossa | 15 | 311,611 | 21,495 | 3 |
|  | Porto Alegre | 163 | 3,581,951 | 19,206 | 8 |
|  | Porto Seguro | 7 | 126,929 | 16,062 | 2 |
|  | Porto Velho | 23 | 448,306 | 18,202 | 16 |
|  | Pouso Alegre | 6 | 130,615 | 18,675 | 1 |
|  | Presidente Prudente | 13 | 231,123 | 15,309 | 1 |
|  | Recife | 116 | 3,513,174 | 29,686 | 3 |
|  | Resende | 8 | 165,144 | 18,708 | 4 |
|  | Ribeirao Preto | 18 | 573,030 | 31,497 | 1 |
|  | Rio Branco | 8 | 336,038 | 42,494 | 1 |
|  | Rio Claro | 11 | 207,887 | 17,868 | 1 |
|  | Rio Grande | 11 | 197,228 | 15,784 | 16 |
|  | Rio Verde | 8 | 176,424 | 18,367 | 3 |
|  | Rio das Ostras | 9 | 141,023 | 13,287 | 2 |
|  | Rio de Janeiro | 332 | 11,737,101 | 27,907 | 2 |
|  | Rondonopolis | 11 | 195,476 | 16,522 | 6 |
|  | Salvador | 88 | 3,200,122 | 29,702 | 2 |
|  | Santa Cruz do Sul | 6 | 118,374 | 17,358 | 4 |
|  | Santa Maria | 16 | 261,031 | 14,006 | 12 |
|  | Santarem | 11 | 294,580 | 28,791 | 1 |
|  | Santos | 57 | 1,569,186 | 24,946 | 8 |
|  | Sao Carlos | 14 | 221,950 | 14,376 | 2 |
|  | Sao Jose dos Campos | 25 | 925,887 | 30,461 | 4 |
|  | Sao Luis | 40 | 1,270,702 | 32,908 | 1 |
|  | Sao Paulo | 618 | 19,356,718 | 30,354 | 6 |
|  | Sertaozinho | 6 | 110,074 | 16,290 | 1 |
|  | Sete Lagoas | 13 | 214,152 | 15,963 | 1 |
|  | Sobral | 9 | 188,233 | 18,143 | 3 |
|  | Sorocaba | 26 | 722,733 | 27,540 | 5 |
|  | Tatui | 4 | 107,326 | 29,668 | 2 |
|  | Taubate | 23 | 466,665 | 16,656 | 1 |
|  | Teresina | 23 | 969,690 | 31,055 | 1 |
|  | Toledo | 7 | 119,313 | 13,813 | 1 |
|  | Tubarao | 8 | 129,544 | 13,938 | 8 |
|  | Uberaba | 16 | 295,988 | 18,179 | 4 |
|  | Uberlandia | 17 | 604,013 | 32,159 | 3 |
|  | Uruguaiana | 6 | 125,435 | 16,676 | 24 |
|  | Varginha | 7 | 123,081 | 17,021 | 2 |
|  | Vitoria | 72 | 1,565,393 | 19,349 | 1 |
|  | Vitoria de Santo Antao | 6 | 129,974 | 23,816 | 2 |
|  | Volta Redonda | 26 | 553,113 | 20,032 | 1 |
| Chile | Antofagasta | 95 | 357,497 | 3,642 | 6 |
|  | Arica | 61 | 164,640 | 2,496 | 4 |
|  | Calama | 45 | 154,693 | 3,328 | 4 |
| Chile | Chillan | 77 | 208,988 | 2,430 | 1 |
|  | Concepcion | 347 | 942,899 | 2,758 | 1 |
|  | Copiapo | 51 | 130,279 | 2,574 | 6 |
|  | Curico | 55 | 143,808 | 2,550 | 1 |
|  | La Serena-Coquimbo | 130 | 413,302 | 3,154 | 1 |
|  | Los Angeles | 64 | 192,720 | 3,018 | 1 |
|  | Osorno | 61 | 161,300 | 2,651 | 1 |
|  | Rancagua | 124 | 325,971 | 2,008 | 1 |
|  | San Antonio | 60 | 126,847 | 1,688 | 1 |
|  | Talca | 106 | 269,130 | 2,178 | 1 |
|  | Temuco | 115 | 357,557 | 2,902 | 1 |
|  | Valdivia | 54 | 159,543 | 3,062 | 1 |
|  | Valparaiso-Vina del Mar | 379 | 940,924 | 2,362 | 1 |
| Colombia | Barrancabermeja | 59 | 157,497 | 2,040 | 10 |
|  | Barranquilla | 196 | 1,732,976 | 7,642 | 11 |
|  | Bogota | 537 | 7,320,947 | 9,351 | 4 |
|  | Bucaramanga | 118 | 996,084 | 6,253 | 1 |
|  | Buenaventura | 77 | 227,879 | 849 | 9 |
|  | Buga | 32 | 110,980 | 3,056 | 6 |
|  | Cali | 293 | 1,627,532 | 3,459 | 1 |
|  | Cartagena | 133 | 905,780 | 5,075 | 6 |
|  | Cartago | 23 | 117,395 | 4,113 | 2 |
|  | Cucuta | 135 | 716,896 | 3,537 | 1 |
|  | Duitama | 19 | 147,834 | 5,767 | 9 |
|  | Girardot | 43 | 122,001 | 1,532 | 1 |
|  | Ibague | 70 | 476,972 | 4,024 | 1 |
|  | Manizales | 86 | 403,658 | 3,260 | 2 |
|  | Medellin | 312 | 3,093,427 | 8,274 | 2 |
|  | Monteria | 63 | 413,332 | 3,659 | 6 |
|  | Neiva | 85 | 313,315 | 2,428 | 5 |
|  | Palmira | 41 | 277,580 | 4,567 | 2 |
|  | Pasto | 39 | 349,346 | 7,862 | 10 |
|  | Pereira | 64 | 650,289 | 9,326 | 3 |
|  | Quibdo | 30 | 119,481 | 268 | 4 |
|  | Riohacha | 72 | 169,941 | 1,532 | 10 |
|  | Santa Marta | 108 | 481,815 | 2,160 | 10 |
|  | Sogamoso | 15 | 118,963 | 8,694 | 1 |
|  | Tulua | 49 | 161,636 | 2,171 | 2 |
|  | Tunja | 28 | 181,513 | 6,204 | 1 |
|  | Valledupar | 89 | 422,067 | 1,337 | 6 |
|  | Villavicencio | 58 | 432,687 | 7,055 | 1 |
|  | Yopal | 33 | 139,029 | 1,672 | 1 |
| Costa Rica | San Jose | 169 | 2,207,560 | 9,727 | 1 |
| Guatemala | Guatemala City | 3,242 | 2,216,383 | 659 | 5 |
| Mexico | Acapulco de Juarez | 506 | 862,727 | 1,315 | 7 |
|  | Acayucan | 56 | 112,922 | 1,156 | 4 |
|  | Acuna | 108 | 136,755 | 1,176 | 7 |
|  | Aguascalientes | 327 | 932,369 | 2,285 | 7 |
|  | Campeche | 121 | 258,412 | 1,818 | 12 |
|  | Cancun | 352 | 676,954 | 1,542 | 12 |
|  | Celaya | 419 | 766,909 | 494 | 4 |
| Mexico | Chetumal | 126 | 243,750 | 1,338 | 12 |
|  | Chihuahua | 639 | 852,527 | 1,227 | 7 |
|  | Chilpancingo | 186 | 241,528 | 672 | 2 |
|  | Ciudad Valles | 107 | 167,643 | 1,128 | 7 |
|  | Ciudad del Carmen | 73 | 221,083 | 2,633 | 12 |
|  | Coatzacoalcos | 135 | 337,696 | 2,396 | 9 |
|  | Cuauhtemoc | 123 | 146,857 | 929 | 1 |
|  | Cuernavaca | 446 | 915,218 | 1,728 | 1 |
|  | Culiacan | 436 | 857,459 | 1,692 | 4 |
|  | Durango | 371 | 572,610 | 1,186 | 3 |
|  | Ensenada | 324 | 464,509 | 1,026 | 5 |
|  | Fresnillo | 172 | 210,428 | 468 | 3 |
|  | Guadalajara | 1,667 | 4,418,566 | 2,458 | 8 |
|  | Guanajuato | 111 | 171,439 | 714 | 7 |
|  | Guaymas | 163 | 198,014 | 962 | 1 |
|  | Hermosillo | 513 | 782,611 | 1,130 | 1 |
|  | Hidalgo del Parral | 75 | 107,015 | 1,462 | 2 |
|  | Iguala | 148 | 140,189 | 419 | 6 |
|  | Irapuato | 163 | 529,399 | 2,305 | 3 |
|  | Juarez | 600 | 1,326,218 | 1,950 | 1 |
|  | La Paz | 211 | 250,509 | 884 | 5 |
|  | La Piedad | 93 | 249,394 | 1,359 | 4 |
|  | Leon | 584 | 1,599,149 | 2,019 | 3 |
|  | Los Mochis | 240 | 416,237 | 1,266 | 4 |
|  | Manzanillo | 123 | 161,420 | 969 | 4 |
|  | Matamoros | 286 | 489,094 | 1,434 | 17 |
|  | Mazatlan | 231 | 436,064 | 1,749 | 4 |
|  | Merida | 557 | 973,031 | 1,590 | 1 |
|  | Mexicali | 459 | 936,616 | 1,658 | 5 |
|  | Mexico City | 5,647 | 20,067,200 | 3,192 | 7 |
|  | Minatitlan | 140 | 356,137 | 2,062 | 9 |
|  | Monclova | 200 | 311,445 | 1,510 | 1 |
|  | Monterrey | 1,610 | 4,104,297 | 2,446 | 6 |
|  | Morelia | 384 | 829,561 | 1,674 | 4 |
|  | Nuevo Laredo | 226 | 384,030 | 1,390 | 2 |
|  | Obregon | 246 | 404,118 | 1,109 | 1 |
|  | Ocotlan | 77 | 140,986 | 1,548 | 8 |
|  | Pachuca de Soto | 352 | 506,556 | 1,096 | 1 |
|  | Piedras Negras | 152 | 178,748 | 1,058 | 6 |
|  | Playa del Carmen | 95 | 159,310 | 998 | 11 |
|  | Poza Rica de Hidalgo | 214 | 513,510 | 1,660 | 10 |
|  | Puebla de Zaragoza | 1,072 | 2,847,343 | 2,394 | 6 |
|  | Puerto Vallarta | 311 | 379,614 | 830 | 4 |
|  | Queretaro | 390 | 1,083,305 | 2,102 | 4 |
|  | Reynosa | 380 | 714,726 | 1,496 | 3 |
|  | Rio Verde | 46 | 135,423 | 1,798 | 1 |
|  | Salamanca | 91 | 260,729 | 1,934 | 2 |
|  | Saltillo | 392 | 822,833 | 1,877 | 2 |
|  | San Francisco del Rincon | 74 | 182,365 | 710 | 3 |
|  | San Juan Bautista Tuxtepec | 56 | 155,752 | 1,866 | 1 |
|  | San Juan del Rio | 92 | 240,175 | 1,937 | 5 |
| Mexico | San Luis Potosi | 380 | 1,033,547 | 2,615 | 4 |
|  | San Luis Rio Colorado | 224 | 178,101 | 829 | 5 |
|  | Santo Domingo Tehuantepec | 112 | 158,765 | 1,215 | 6 |
|  | Tampico | 402 | 855,026 | 1,922 | 10 |
|  | Tapachula | 162 | 320,173 | 1,308 | 7 |
|  | Tecoman | 99 | 140,600 | 1,286 | 4 |
|  | Tehuacan | 125 | 296,681 | 2,165 | 1 |
|  | Tepic | 204 | 429,302 | 1,832 | 8 |
|  | Tianguistenco | 57 | 157,944 | 1,811 | 5 |
|  | Tijuana | 654 | 1,675,065 | 2,307 | 5 |
|  | Tlaxcala | 300 | 499,551 | 1,369 | 1 |
|  | Toluca | 563 | 2,012,208 | 2,755 | 6 |
|  | Torreon | 663 | 1,214,063 | 1,519 | 2 |
|  | Tula de Allende | 160 | 286,398 | 1,253 | 7 |
|  | Tulancingo de Bravo | 121 | 236,893 | 1,428 | 6 |
|  | Tuxtla Gutierrez | 260 | 681,944 | 2,348 | 6 |
|  | Uriangato | 113 | 108,455 | 105 | 3 |
|  | Uruapan | 159 | 312,610 | 1,620 | 4 |
|  | Veracruz | 363 | 810,298 | 2,006 | 12 |
|  | Victoria | 176 | 321,953 | 1,602 | 1 |
|  | Villahermosa | 177 | 755,425 | 3,012 | 9 |
|  | Zacatecas | 288 | 307,260 | 794 | 2 |
|  | Zamora | 116 | 250,091 | 1,884 | 2 |
| Panama | Colon | 158 | 199,993 | 563 | 1 |
|  | David | 298 | 196,399 | 296 | 6 |
|  | Panama City | 1,062 | 1,379,040 | 560 | 7 |

**Supplemental Table S3:** Multilevel associations between flooding and urban features comparing main analysis (including all cities with one or more recorded flood events) and sensitivity analysis (including only cities with at least two recorded flood events).

|  |  | Main analysis: 1+ flood events  (N=276 cities) | | Sensitivity analysis:  2+ flood events  (N=196 cities) | |
| --- | --- | --- | --- | --- | --- |
|  |  | Estimate | 95% CI | Estimate | 95% CI |
|  | Population density* | **0.47** | 0.41, 0.52 | **0.46** | 0.40, 0.52 |
|  | Education | **0.81** | 0.76, 0.86 | **0.81** | 0.75, 0.87 |
|  | Intersection density | **0.71** | 0.65, 0.78 | **0.78** | 0.71, 0.85 |
| Neighborhood- | Greenness | **1.43** | 1.32, 1.55 | **1.52** | 1.40, 1.66 |
| Level | Distance from city center | **1.19** | 1.13, 1.25 | **1.16** | 1.10, 1.23 |
|  | Coastal (<1 km) | **46.6** | 38.6, 56.3 | **62.4** | 49.9, 78.0 |
|  | Altitude | **1.39** | 1.16, 1.66 | **1.59** | 1.29, 1.96 |
|  | Slope | **0.79** | 0.75, 0.84 | **0.76** | 0.71, 0.82 |
|  | Population size | 0.95 | 0.83, 1.09 | 0.92 | 0.77, 1.10 |
|  | Population density | 0.93 | 0.72, 1.19 | 0.84 | 0.62, 1.15 |
|  | Education | 0.95 | 0.72, 1.25 | 0.97 | 0.71, 1.32 |
| City-level | Intersection density | 0.99 | 0.83, 1.19 | 0.97 | 0.78, 1.20 |
|  | Greenness/vegetation | 0.97 | 0.78, 1.20 | 0.87 | 0.66, 1.15 |
|  | GDP | 1.03 | 0.87, 1.20 | 1.05 | 0.86, 1.28 |
|  | Climate |  |  |  |  |
|  | Temperate | 1 (Ref.) |  | Ref |  |
|  | Arid | 0.62 | 0.36, 1.07 | 0.61 | 0.32, 1.17 |
|  | Tropical | 1.15 | 0.78, 1.70 | 1.18 | 0.75, 1.88 |

All independent variables have been z-transformed using the distribution of all study neighborhoods, except for proximity to coast which is binary (yes/no <1 km from coast) and climate zone (categorical). Estimates in bold font represent estimates with statistical significance of p<0.05.

**Supplemental Figure S1**: Spearman correlation coefficients for covariates in the main model of the associations between neighborhood flooding and features of the urban environment.

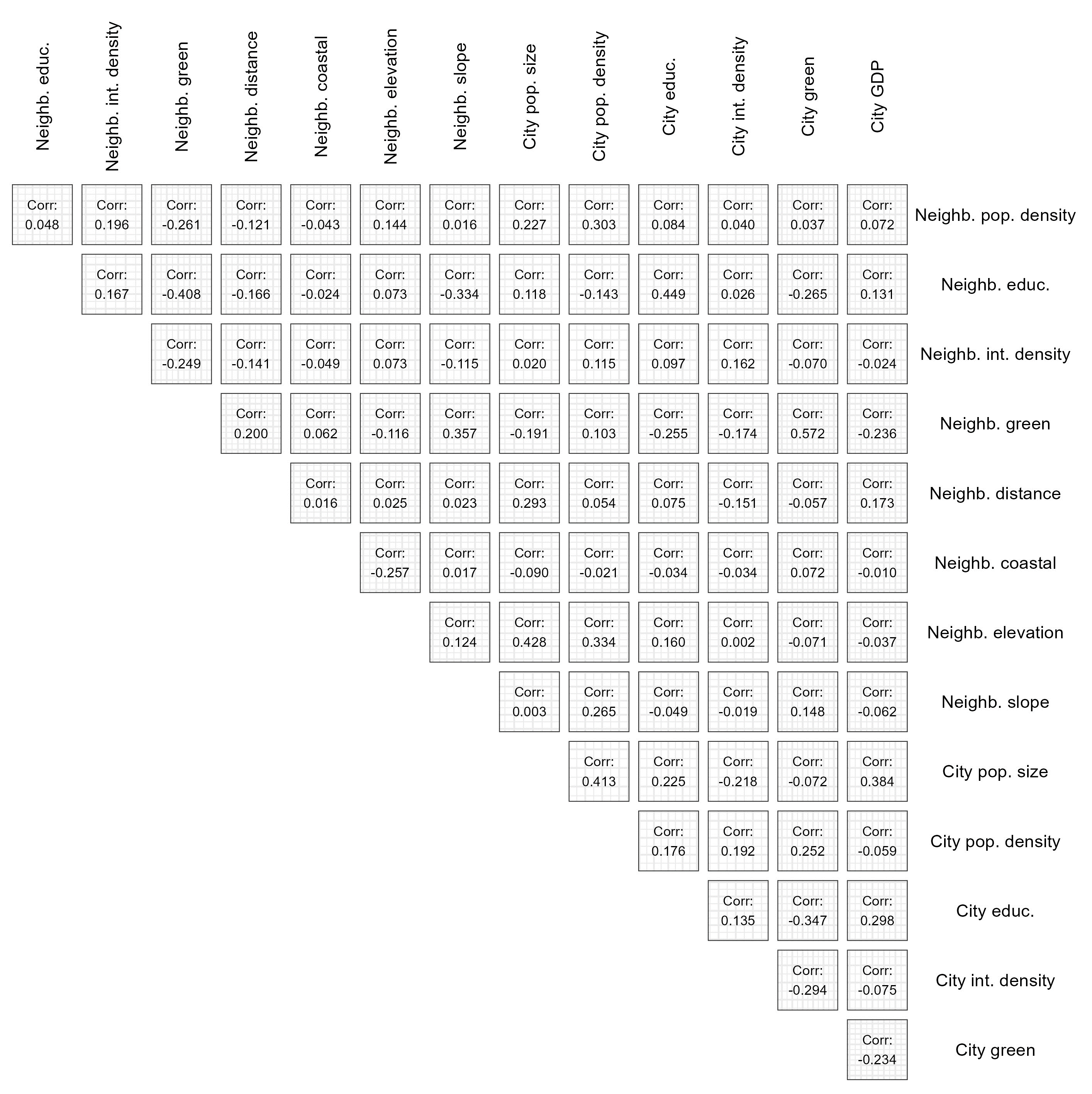

**Supplemental Figure S2**: Within-city variation in neighborhood flooding in two selected cities: (a) Buenos Aires, Argentina and (b) Medellin, Colombia. Black lines represent neighborhood boundaries and colors represent number of distinct floods from 2000-2018 in the Global Flood Database. The metropolitan area of Buenos Aires, Argentina (left panel), and Medellín, Colombia (right panel).

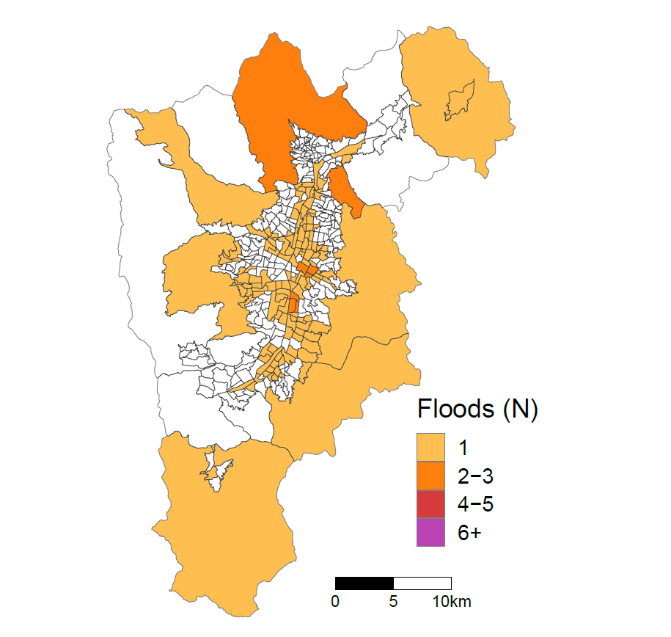

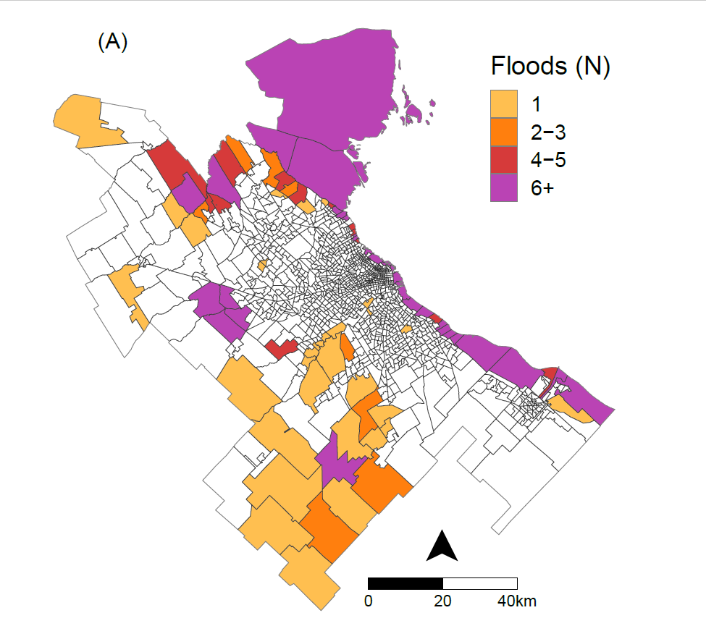

**Appendix I**: Detailed descriptions of urban environmental measures and their interpretation

**Population** (neighborhood/city). Number of residents reported by most recent available national census (see Supplemental Table S1 for census year for each country).

**Population Density** (neighborhood/city). Population (in thousands) per square kilometer of neighborhood/city area. Population sourced from national censuses.

**Educational attainment** (neighborhood/city). Percent of the population aged 25 years or older who completed primary education or above. Sourced from national censuses.

**Intersection Density** (neighborhood/city). The number of intersections per area of city/neighborhood built-up area in square kilometers. Intersections were extracted from street network OpenStreetMap data and included any intersections with >2 connected streets (i.e., cul-de-sacs and road bends represented as a node were excluded). Neighborhood and city indices represent the years of 2020 and 2017, respectively.

**Greenness** (neighborhood/city). Area median greenness measured by the normalized difference vegetation index (NDVI). NDVI was calculated using MODIS satellite-based observations from the MODIS vegetation product, MOD13Q1.006 for 2015 at a 250 m spatial resolution. We computed the maximum NDVI value for 2019 at 250 m resolution to present the ‘greenest’ condition of each grid cell within the year, then calculated the median across grid cells contained within each neighborhood.

**Distance from city center** (neighborhood). Euclidean distance in km between the neighborhood centroid and city hall, based on neighborhood boundaries at time of census.

**Gross Domestic Product per Capita** (city). City-level GDP in 2011 international US Dollars. Created by Genaioli et al in 2013 and converted into gridded estimates by Kummu et al in 2015. GDP for each year 1990-2015 was estimated by these researchers by modeling data from government, survey and industry. Gridded estimates were matched to SALURBAL cities and GDP was extracted directly from matching administrative units or using population-weighted averages in cases where city boundaries crossed multiple administrative areas. GDP estimates from 2015 (most recent year available) were used.

Gennaioli N, La Porta R, Lopez-de-Silanes F, Shleifer A, Human Capital and Regional Development, The Quarterly Journal of Economics, Volume 128, Issue 1, February 2013, Pages 105–164, https://doi.org/10.1093/qje/qjs050.

Kummu M, Taka M, & Guillaume J. Gridded global datasets for Gross Domestic Product and Human Development Index over 1990–2015. Sci Data 5, 180004 (2018). https://doi.org/10.1038/sdata.2018.4.

**Coastal Neighborhood.** Coastal geographies are those with parts of their boundaries within 1 km from the closest ocean coastline, whereas inland regions represent the rest**.** Coastal lines are obtained from Global Self-consistent, Hierarchical, High-resolution Geography Database (GSHHG) (https://www.ngdc.noaa.gov/mgg/shorelines/, high resolution version).
